# Tremor Morphology Toolkit: Waveform Modulation-Based Tremor Phenotyping

**DOI:** 10.64898/2026.11.05.26367334

**Authors:** Prajakta Joshi, Yuri Ferreira Felloni Borges, Sanjay Pandey, Marie Vidailhet, Victor Fung, Mark Hallett, Hyder A. Jinnah, Alfanso Fasano, Aasef G. Shaikh

## Abstract

Clinicians typically characterize tremor by its frequency, amplitude, and body region affected. Other aspects also provide critical clues to the type of tremor and its underlying pathophysiology including tremor waveform morphology, consistency across cycles, and variability in amplitude and timing. We introduce the Tremor Morphology Toolkit (TMT), an eight-metric, waveform-derived panel that quantifies these features from upper-limb inertial signals. Using accelerometer and gyroscope recordings from 34 individuals with upper-extremity tremor across four standardized tasks (rest, outstretched, wing-beating, and kinetic), we computed: (i) rSHAPE, cycle-to-cycle shape consistency via pairwise similarity of normalized cycles; (ii) rSINE, sinusoidality, the mean correlation of each cycle with its best-fit sinusoid; (iii) rCVA, the coefficient of variation of cycle amplitude; (iv) rCVD, the coefficient of variation of cycle duration; (v) AWI, amplitude-warping index; (vi) DWI, duration-warping index; (vii) FreqVar, a spectral bandwidth indexing power-weighted frequency spread; and (viii) Amplitude Breathing indices, which quantify slow waxing and waning of tremor amplitude over time from fluctuations in the signal envelope, representing the time-varying amplitude of the tremor waveform. We detail algorithms, parameterizations, and implementation defaults to support reproducible use. TMT is designed to complement conventional frequency–amplitude measures by capturing tremor morphology and modulation in a simple, interpretable framework. It enables objective quantification of features often assessed qualitatively at the bedside, facilitates comparisons across recording conditions, and provides a foundation for validation against clinical labels and disease phenotypes. Future studies can integrate these parameters with electrophysiological measures to investigate mechanisms underlying tremor generation and modulation.

**Highlights:**

1. Introducing Tremor Morphology Toolkit (TMT), an eight-metric framework for objective tremor characterization.
2. Correlation analysis identified redundant and complementary relationships among tremor-shape features, with sinusoidal waveform characteristics (rSINE), cycle-to-cycle amplitude variability (rCVA), and tremor frequency variability (FreqVar) providing a concise, complementary set for characterizing tremor shape.
3. Examination of the alignment between newly defined tremor-shape features and clinician perceptions found that clinicians relied more on envelope-based amplitude dynamics than on individual tremor cycles.

## Introduction

“Tremor” is often used as an umbrella term for repetitive movements with multiple subtypes, each with distinct pathophysiology making precise characterization crucial for diagnosis and treatment. It is defined as an involuntary, oscillatory, repetitive back-and-forth movement of a body part about a fixed point, axis, or plane (Crawford and Zimmerman, 2018; Shaikh et al., 2024)). Traditionally, tremor is described by its frequency, amplitude, distribution (location, symmetry and laterality), and activation (rest vs action) (Albanese and Sorbo, 2016; Algarni and Fasano, 2018; Alty and Kempster, 2011; Stouwe et al., 2020; Thenganatt and Jankovic, 2016). Beyond rate and magnitude, the trajectory shape i.e. morphology of the movement can provide additional diagnostic cues. Clinicians visually assess whether the path is regular or jerky and how consistent that pattern remains over time. Qualitative descriptors that complement these metrics include amplitude variability, frequency variability, the shape of the movement trajectory, the consistency of that shape over time. Several of these features can be subtle or require extensive expertise to detect, allowing subjective weighting of parameters to influence the assessment. Such limitations contribute to frequent misdiagnosis, delaying optimal therapy (Rizzo et al., 2016).

Imaging and deep brain stimulation studies link essential tremor–like head oscillations to cerebellar circuits projecting via the ventral intermediate (VIM) thalamus and dentatothalamic tracts, which clinically manifest as regular, rhythmic cycles (Erro et al., 2022; Panyakaew et al., 2022). In contrast, oscillations co-occurring with dystonia are characterized by impaired inhibition at multiple levels and pallidal unit activity with burst–pause patterns, yielding irregular, “saw-tooth” kinematics and frequent antagonist co-contraction or overflow on EMG (Ulia Semenova et al., 2021). Cerebellar-receiving thalamic firing differed by phenotype: patients with pure jerky head oscillations showed predominantly single-spike activity, whereas those with dystonia plus jerky oscillations exhibited a more bursting/mixed pattern. In contrast, pallidal-receiving thalamic nuclei showed no phenotype-dependent shift, supporting a pathophysiological basis for differences in movement-shape manifestations (U. Semenova et al., 2021). Converging network analyses and DBS outcomes further indicate that many patients exhibit combined cerebello-thalamo-cortical (CTC) and basal ganglia–thalamo-cortical (BGTC) contributions (Albanese and Sorbo, 2016; Algarni and Fasano, 2018; Erro et al., 2022; Karachi et al., 2005; Logroscino et al., 2003; Shaikh et al., 2024). Consistent with these clinical–physiological distinctions, spectral measures such as half-power bandwidth (Murgai and Iskhakova, 2025) and the Tremor Stability Index (Biase et al., 2017) tend to broaden and destabilize with jerky morphology, aiding discrimination of dystonia when present with tremor from the more regular ET-like pattern. Hence, metrics that address waveform shape and its variance over time are needed to complement frequency and amplitude, capturing regularity, cycle morphology, and modulation stability that map onto underlying circuit contributions.

Traditional tremor characterization has primarily emphasized the body distribution of tremor, along with its amplitude and frequency. Clinical rating scales such as the Tremor Research Group Essential Tremor Rating Assessment Scale (TETRAS) similarly place substantial emphasis on tremor amplitude across different body regions and tasks (Elble, 2016). However, tremors with similar amplitudes or dominant frequencies may differ in their waveform structure and temporal behavior, suggesting that additional characteristics including waveform shape, amplitude dynamics, and cycle-to-cycle frequency stability may provide complementary information about tremor phenomenology.

The definitions of these features can be subjective and prone to varying interpretations. Our interrater reliability study using a basic trajectory shape revealed markedly lower agreement for tremor morphology than for broader characteristics such as presence, symmetry, and frequency, underscoring the need for an objective, standardized framework (Joshi et al., 2026). It also exposed confounding between amplitude and frequency variability - each can bias interpretation of the other. Additionally, it showed that subjective assessment may misinterpret constant amplitude waveform as a sinusoidal waveform. Whether these discrepancies reflect true physiology or clinician bias, they motivate methods that (i) disentangle frequency modulation from amplitude-based modulation and (ii) isolate amplitude effects from waveform shape. We therefore introduce the Tremor Morphology Toolkit (TMT) with eight features, an objective framework that independently quantifies amplitude variability, frequency variability, waveform-shape consistency, and sinusoidal nature (how closely the average cycle of tremor approximates a sine wave) from inertial-measurement-unit accelerometer and gyroscope data suited to the non-stationary nature of tremor signals.

## Methods

### Participants and Ethical Approval

This study was performed in accordance with the Declaration of Helsinki and approved by the University of Toronto Institutional Review Board. All participants provided written informed consent prior to enrollment. Data were collected from 34 participants with clinically diagnosed upper-limb tremor. Based on clinical assessment, participants were categorized as having essential tremor (n = 16), Parkinsonian tremor (n = 6), dystonic tremor (n = 4), or other tremor (n = 8). Tremor was assessed during standard positions and tasks, including rest (arms resting on the armchair), postural tremor (arms extended at shoulder height), wing tremor (forearms folded toward the chest with elbows lifted outward), and kinetic tremor (index-to-nose maneuver).

### Data Acquisition

Tremor recordings were obtained using an Xsens DOT inertial measurement unit (IMU; Movella Inc., Enschede, The Netherlands) secured to the participant’s palm with an adjustable strap. Data were sampled at 60 Hz for a minimum of 30 s per trial. Each recording was stored in .xlsx format for subsequent analysis. A videotape was also obtained as the participants performed the tasks and subsequently rated by four movement disorder neurologists using the Tremor Waveform Morphology Questionnaire (Joshi et al., 2026). The raters were blinded to the participants’ clinical diagnoses.

### Signal Preprocessing

All analyses were performed in MATLAB. We used the Madgwick orientation filter (Madgwick, 2010) with β = 0.01 to fuse accelerometer, gyroscope, and magnetometer data and estimate roll, pitch, and yaw angles. We then formed a composite position vector (CPV) by taking the root-mean-square (RMS) of the three angles, and band-pass filtered the CPV (1–15 Hz; 4th-order Butterworth) to isolate the oscillatory component (OC) of interest as illustrated in Figure 1.

**Figure 1.**
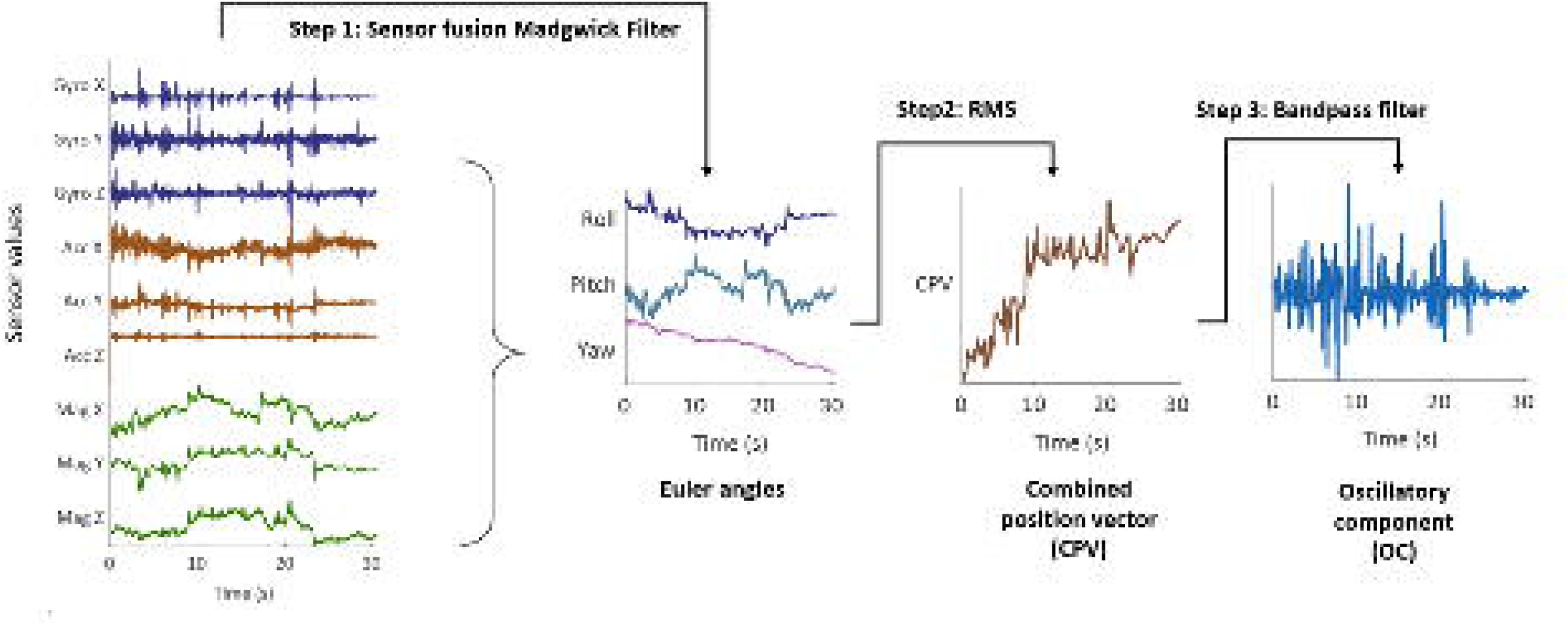
Computation of the oscillatory component (OC) from inertial sensor data: Accelerometer, gyroscope, and magnetometer signals were fused using the Madgwick orientation filter (Madgwick, 2010; β = 0.01) to estimate roll, pitch, and yaw angles. A composite position vector (CPV) was then obtained as the root-mean-square (RMS) of the three orientation angles. The CPV was subsequently band-pass filtered (1–15 Hz; 4th-order Butterworth) to extract the oscillatory component of interest.

**Figure 2:**
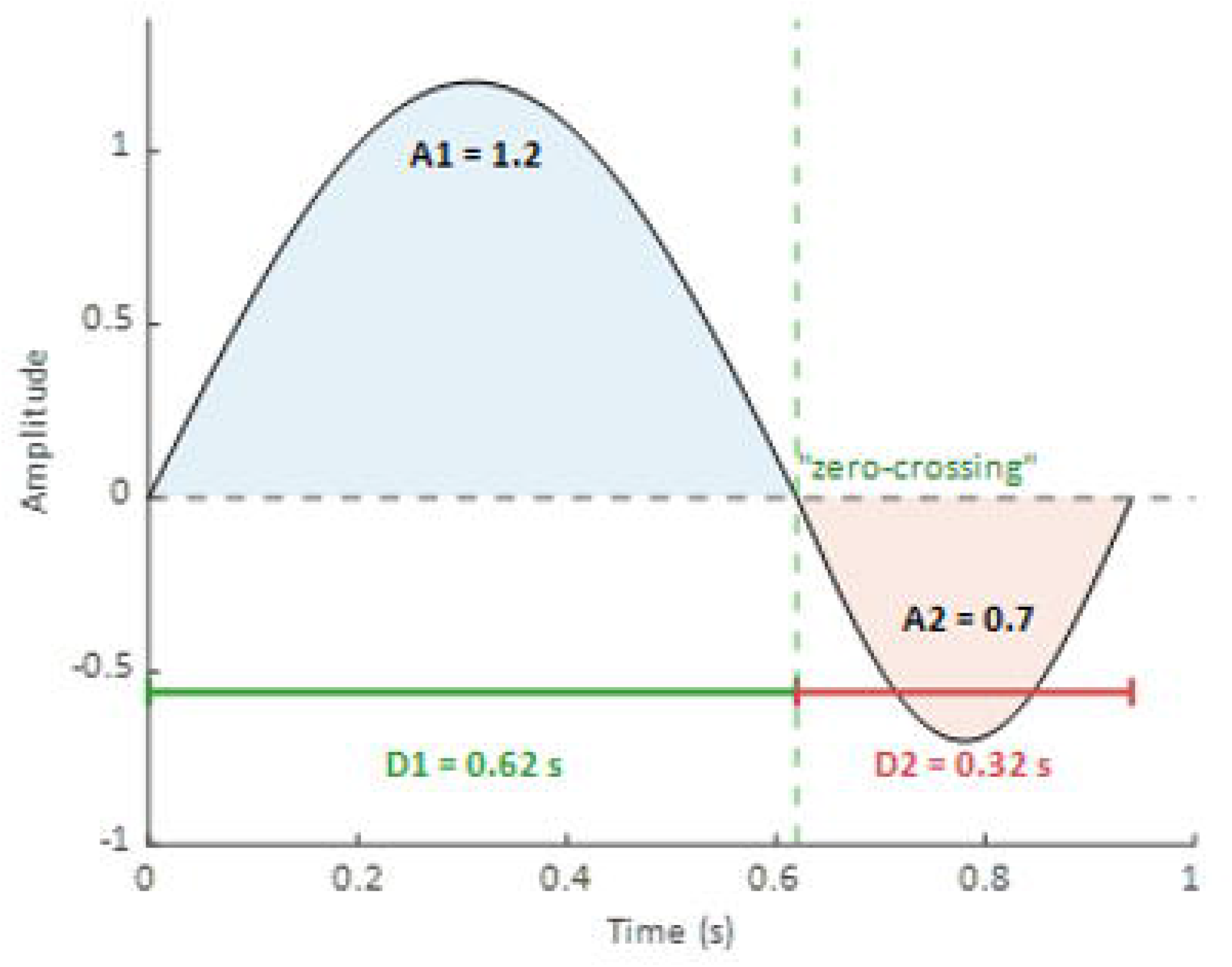
Example of a “cycle”. A1, A2 are the amplitudes of positive and negative lobe, and D1, D2 are the durations.

### Tremor Morphology Toolkit description

In the time domain, we identified zero-crossings and defined a “cycle” as the interval between successive alternate zero-crossings. Each new cycle therefore begins at every other zero-crossing. The first six indices were derived by summarizing features extracted from individual cycles: Amplitude Modulation Index, Frequency Modulation index, Sinusoidality Index, Shape Similarity Index, Amplitude Warping Index, and Cycle Duration warping index. The next measure describes the frequency variability which account for the power at various frequencies in the spectrum. The last one is a set of amplitude breathing indices which attempt to quantify the waxing and waning patterns of amplitude change in an oscillatory tremor like movement.

1. *<u>Amplitude Modulation Index (rCVA)</u>*: For each cycle, the peak-to-peak amplitude was computed as the difference between the maximum and minimum values. Variability across cycles was then quantified using a robust coefficient of variation (rCVA) computed over the distribution of amplitudes given by equation (1). A is an array of per-cycle amplitudes and MAD is the median absolute deviation of A.

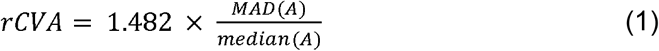 We use a robust statistic because cycle amplitudes in tremor data are may not be Gaussian: motion artifacts, brief co-contractions, sensor clipping, and missed segments can produce outliers, skew, and heavy tails. The standard CV (standard deviation/mean) is highly sensitive to such extremes and can overstate variability. In contrast, the MAD/median formulation has a 50% breakdown point, greatly reducing the influence of outliers while still reflecting the distribution’s spread; scaling MAD by 1.4826 makes it comparable to SD under normality (Huber and Ronchetti, 2023). This yields stable, comparable variability estimates across tasks and subjects, even when a few cycles are aberrant.
2. <u>Cycle-duration Modulation index (rCVD)</u>: We used cycle duration as the time-domain proxy for frequency. It is defined as the interval between successive same-phase zero-crossings (i.e., one period). Analogous to rCVA, variability in cycle duration across cycles was summarized with a robust coefficient of variation to yield the frequency modulation index - rCVD using equation (2).

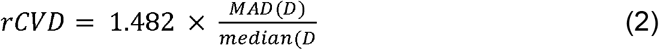 D is an array of per-cycle amplitudes and MAD is the median absolute deviation of D.
3. *<u>Sinusoidality Index (rSINE)</u>*: To quantify how closely each waveform cycle resembles a sine wave, we defined the sinusoidality index (rSINE). All cycles were time-normalized to 100 samples by interpolation. We averaged these to obtain a mean cycle and generated a reference sine wave matched to its amplitude. For each cycle, we computed the absolute Pearson correlation with this reference; the mean of these correlations across cycles is rSINE. Higher rSINE indicates a more sinusoidal cycle shape.
4. *<u>Shape Similarity Index (rSHAPE)</u>*: rSHAPE quantifies cycle-to-cycle waveform similarity. All cycles were time-normalized to 100 samples by interpolation and averaged to form a mean template. For each cycle, we computed the absolute Pearson correlation with this template; the mean of these correlations across cycles defines rSHAPE (higher values = closer match to the template).
5. <u>Amplitude Warping Index (AWI)</u>: The “Amplitude Warping Index” (AWI) captures intra-cycle amplitude asymmetry between a waveform’s two lobes (e.g., positive vs. negative half-cycles). It reports how unequal the lobe amplitudes are, independent of overall signal scale. We define,

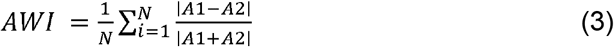

where N is the total number of cycles in the recorded signal, and A1 and A2 are amplitudes of each lobe. 0 indicates perfectly matched lobe amplitudes, larger values indicate greater amplitude asymmetry.
6. <u>Duration Warping Index (DWI)</u>: The “Duration warping Index” captures asymmetry in time between the two lobes (positive vs negative half-cycle; D1 and D2) of a waveform cycle. It reflects whether one lobe is stretched (longer) while the other is compressed (shorter). We define

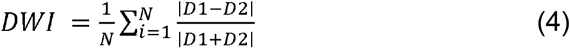

where N is the total number of cycles in the recorded signal. It measures lobe-duration asymmetry (0 = perfectly balanced; larger values = greater distortion).
7. <u>Frequency variability (FreqVar)</u>: We estimated frequency variability (FreqVar) as the bandwidth of the central 66% of spectral energy. For each signal, a continuous wavelet transform (32 voices/octave) was computed; time–frequency power was averaged over time to obtain a mean spectrum, normalized, and its cumulative distribution used to locate the 17th and 83rd percentiles. A larger FreqVar indicates broader spectral spread, while smaller values correspond to more spectrally stable, periodic signals.
8. <u>Amplitude Breathing Indices (ApEn, Tremor Envelope Breathing Rate, Tremor Envelope Modulation Ratio):</u> We quantified “breathing-like” amplitude modulations in the oscillatory component to capture slow waxing-waning of tremor amplitude. These envelope features probe non-stationarity and organization of the signal: (i) the Tremor Envelope Breathing Rate (TEBR) (peaks of the smoothed amplitude envelope per second) indexes how often slow amplitude swells occur; (ii) the Tremor Envelope Modulation Ratio (median peak-to-trough amplitude) summarizes the strength of each swell; and (iii) approximate entropy to measure the unpredictability of these modulations. We quantified these slow “breathing-like” amplitude modulations of the oscillatory component as follows. Upper and lower envelopes were obtained with MATLAB’s peak-based envelope function at two scales: First, a short window (1 sample) was used to calculate the upper and lower envelopes (dark red lines in Figure 3 plots). Instantaneous amplitudes were computed as the difference between upper and lower envelopes. This amplitude series was used to compute approximate entropy (ApEn) as a measure of regularity (lower ApEn = more stereotyped modulation, higher ApEn = more erratic changes). A second envelope was calculated using a larger window (30 samples or half the sampling rate) to capture slower modulations (dark blue line in figure 3 plots). Peaks of the smoothed amplitude envelope defined candidate modulation cycles, and troughs were identified with local minima shown (Figure 3, peaks shown as dark-gray filled circles; troughs as light-gray circles). To summarize modulation depth, we calculated the median peak-to-trough amplitude ratio across aligned cycles (starting from a trough). The TEBR (Hz) was defined as the number of amplitude-envelope peaks divided by the total recording duration, yielding cycles per second of slow amplitude “waxing–waning.” Figure 3 shows four example oscillatory patterns and their calculated indices.

**Figure 3.**
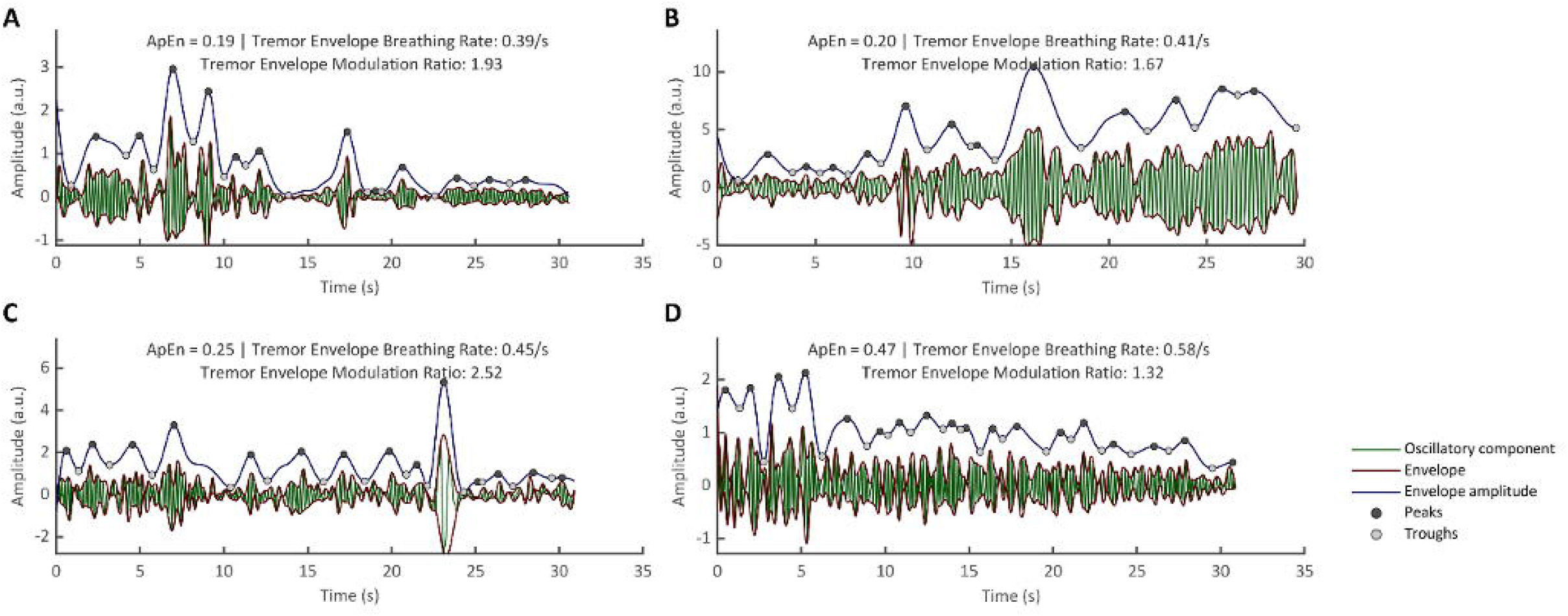
Examples of amplitude “breathing” indices across four tremor signals: Maroon lines depict the envelope; the dark-blue trace is the envelope amplitude (upper–lower envelope); filled dots mark peaks and troughs of the envelope-amplitude series used for index computation. For each panel, we report (i) Breathing Rate (peaks·s⁻¹), (ii) Breath Modulation Ratio: median peak-to-trough ratio, and (iii) Approximate Entropy (ApEn) of the fine-scale envelope amplitude. (A) Regular, highly predictable modulation (ApEn = 0.19), with a median Breathing Rate of 0.39 s⁻¹ and BMI = 1.93. (B) Slower, shallower modulations than (A), reflected in a lower BMI. (C) Larger modulation depth and faster rate, yielding a high BMI and elevated Breathing Rate. (D) Less predictable envelope-amplitude dynamics (ApEn = 0.47) with a relatively high Breathing Rate compared to the other examples.

### Feature Dependence of Clinician Labels

Clinicians classified the observed oscillatory movements using the TWMQ (Joshi et al., 2026). Following prior approaches (reference pending), rater perceptions of tremor presence, amplitude stability (constant or varying), rhythm regularity (regular or irregular), jerkiness (present or absent), and frequency range were modeled using newly defined features in the Tremor Morphology Toolkit. We examined how objectively quantified tremor features correspond to clinicians’ categorical ratings using a supervised learning framework. Each trial (participant × arm position: rest, wing, or kinetic) was represented by eight features: *rCVA*, *rSINE*, *FreqVar*, *peakF*, *peakAmp, TEBR, TEMR, and ApEn*. *peakAmp* and *peakF* are the peak amplitude and the corresponding frequency of the power spectral density curve of the oscillatory component. Trials lacking rater annotations were excluded, and when multiple raters were available, feature rows were replicated to align with individual ratings. For each rating type (tremor presence, amplitude constancy, rhythm, jerkiness, and frequency range), features were z-scored and evaluated using five-fold stratified cross-validation with 10 repetitions, in which each fold comprised stratified training (80%) and test (20%) subsets to preserve class balance. A bagged decision-tree ensemble (*fitcensemble*; 200 trees; minimum leaf size = 10) was trained on the training data, and model performance on the held-out test set was summarized using accuracy and the area under the receiver operating characteristic curve (AUROC). Feature importance was estimated through permutation analysis (10 randomizations per feature), and mean AUROC reductions were normalized to yield percent influence.

## Results

We computed all indices for 136 signals from 34 participants across four conditions: rest, arms outstretched, wing posture, and kinetic movement. Figure 4 illustrates four example tremor movements recorded using the IMU, followed by a pictorial representation of the key processing step used to derive the tremor features included in the Tremor Morphology Toolbox. The first column shows the oscillatory component; the second column displays the power spectral density, which is used to calculate the frequency variability. The third column presents shape-correlation matrices (rSINE, rSHAPE). The fourth and fifth columns contain histograms of cycle amplitudes and cycle durations with the corresponding rCVA and rCVD metrics. The final column overlays all detected cycles to visualize time- and amplitude-warping, together with AWI and DWI. Figure 4A depicts a highly sinusoidal signal with strong shape consistency (high rSINE/rSHAPE), low amplitude variability, minimal cycle-duration variability consistent with low frequency variability, and small warping indices indicating near-symmetric cycles. In contrast, Figure 4B shows marked amplitude variability (higher rCVA than 4A) but relatively low frequency variability, accompanied by elevated AWI and DWI. Figure 3C illustrates a less sinusoidal morphology (rSINE = 0.77) with high amplitude variability and moderate cycle-duration variability (rCVD), together with increased frequency variability. Finally, Figure 4D exhibits pronounced frequency variability with irregular amplitude and cycle-duration patterns, as reflected in the associated metrics.

**Figure 4.**
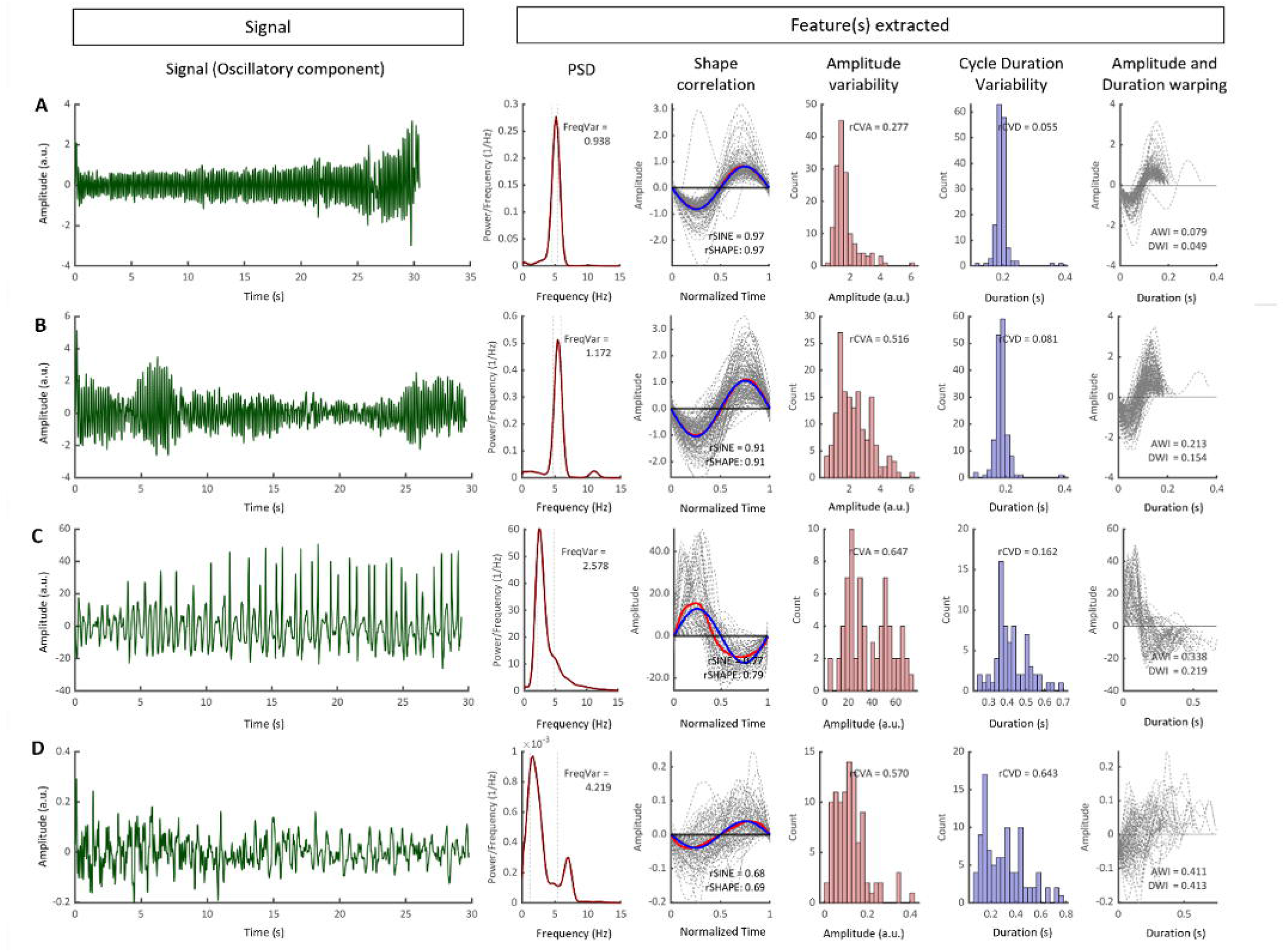
Representative examples illustrating tremor signal morphology features: Each row represents a recording with distinct waveform characteristics. The first column shows the oscillatory component of the signal, and the second column shows its power spectral density (PSD), used to quantify frequency variability. The third column presents the extracted tremor cycles after time normalization, which are used to calculate shape-correlation matrices (rSINE, rSHAPE) quantifying waveform similarity and sinusoidality. The fourth and fifth columns show histograms of cycle amplitudes and durations, with the corresponding rCVA and rCVD indices quantifying amplitude and duration variability, respectively. The final column overlays all extracted cycles to illustrate the extent of time and amplitude warping, quantified by the AWI and DWI indices. Together, these panels demonstrate differences in waveform symmetry, regularity, and frequency stability across representative tremor signals.

### Distribution and interrelationships among tremor-morphology features

We next assessed the distributional profiles of the first seven of newly defined tremor-morphology features. Figure 5A displays histograms for all features across 136 signals, with vertical markers for the mean and median of each distribution. Correlation-based shape metrics lie predominantly between ∼0.60 and 1.00, rCVA and rCVD span ∼0 to 1.20, and the warping indices (AWI, DWI) fall within ∼0 to 0.50. We also tested whether these feature values depended on posture or movement. By conducting one-way ANOVA for each feature across rest, arms-outstretched, wing, and kinetic conditions; no significant effects were detected, indicating that postural context did not influence the values.

**Figure 5.**
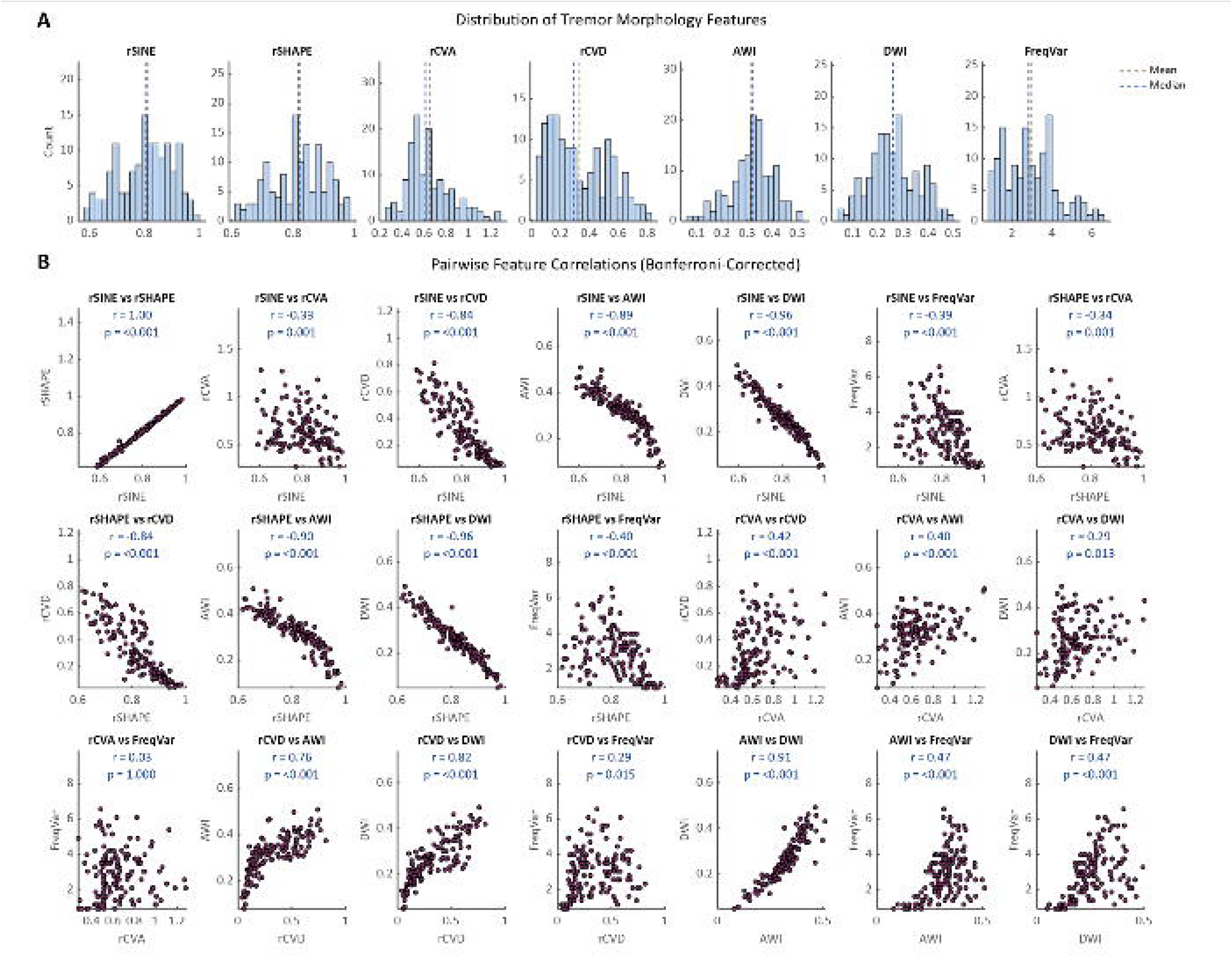
Distribution and interrelationships among tremor-morphology features. (A) Histograms showing the distribution of each feature across 136 oscillatory signals, with vertical lines marking the mean and median. (B) Pairwise correlation matrix (Bonferroni-corrected) illustrating relationships among features.

To identify features that conveyed largely non-overlapping information, we computed pairwise Spearman correlations across all features (Figure 5B). To assess the independence of these features, we used an independence criterion of ∣r∣<0.50; pairs with ∣r∣≥0.50 were considered redundant. Depending on the context and definition it may reduce the amount of new information being conveyed by these parameters. From the first panel of Figure 4B, rSINE and rSHAPE are highly correlated, indicating strong concordance between the two shape-based similarity metrics. Both rSINE and rSHAPE show inverse relationships with cycle-duration variability and with the warping indices (AWI, DWI). Cycle duration variability is itself strongly correlated with both AWI and DWI, and AWI–DWI are tightly coupled (r = 0.91), suggesting that amplitude and duration warping tend to co-occur. FreqVar and rCVA show little correlation with other indices. Taken together, for these 136 tremors, the trio of rSINE, rCVA, and FreqVar provides a concise, complementary set for describing tremor morphology.

### Classification of oscillatory signals by waveform shape and modulation type

We next characterized tremors using the proposed features, aiming to classify each signal as sinusoidal vs. non-sinusoidal, amplitude-constant vs. amplitude-modulated, and frequency-constant vs. frequency-variable. In the absence of external references, we adopted empirical thresholds: rSINE ≥ 0.80 for sinusoidal morphology, rCVA ≥ 0.20 for amplitude modulation, and FreqVar ≥ 2 Hz for frequency variability. Signals not meeting these criteria were labeled non-sinusoidal, amplitude-constant, or frequency-constant, respectively. All 136 signals exceeded the amplitude-modulation threshold (rCVA > 0.20). Table 1 summarizes the distribution: ∼55% (74/136) met the sinusoidal criterion; among these, 31/74 (∼40%) were amplitude-modulated only (AM), while ∼60% showed both amplitude and frequency modulation (AM+FM). Among non-sinusoidal signals, ∼17% were AM-only, with the majority classified as AM+FM.

**Table 1:** Classification of oscillatory signals by waveform shape and modulation type. Counts (with percentages of the full dataset, *N* = 136) are shown for sinusoidal (rSINE ≥ 0.80) vs. non-sinusoidal signals, stratified by modulation: amplitude-modulated (AM; rCVA ≥ 0.20) and amplitude + frequency-modulated (AM+FM; rCVA ≥ 0.20 and frequency variability ≥ 2 Hz). Row and column totals are provided for reference.

|  | Amplitude Modulated (AM) | Amplitude + Frequency modulated (AM + FM) | Total |
|---|---|---|---|
| Sinusoidal | 31 (23%) | 43 (32%) | 74 (55%) |
| Non-sinusoidal | 11 (8%) | 51 (38%) | 62 (45%) |
| Total | 42 (30%) | 94 (70%) | 136 |

### Distribution of Amplitude Breathing

Across the dataset, Amplitude Breathing Indices showed bounded variability (Figure 6). Approximate entropy ranged from 0 to 0.62, Breathing Rate from 0.33 to 0.63 Hz, and the Breath Modulation Ratio from 0.8 to 10. Pairwise associations among these three measures were not significant, and none showed significant relationships with previously computed morphology indices, indicating that the breathing metrics capture largely independent aspects of amplitude dynamics (not shown).

**Figure 6:**
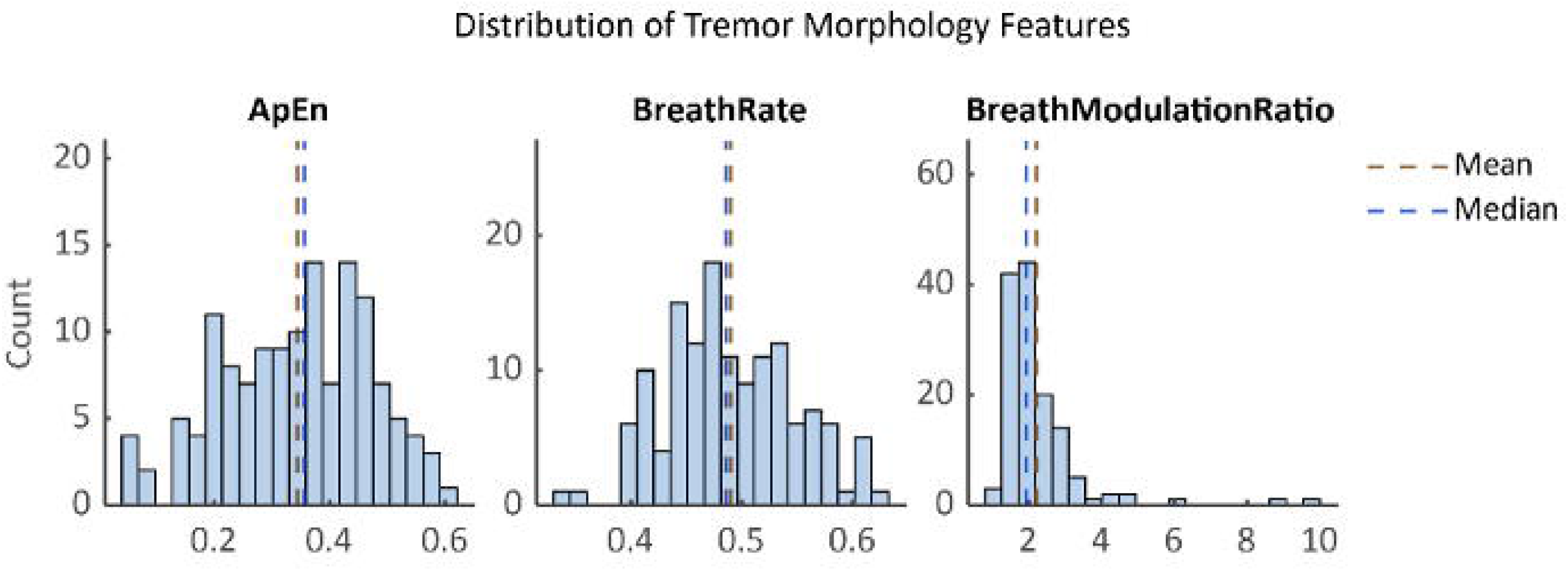
Shows distribution of Amplitude Breathing Indices across the dataset.

### Linking Rater Perception to Quantitative Tremor Morphology Metrics

Figure 7 illustrates the estimated percentage contribution of each independent feature from the Tremor Morphology Toolkit to the corresponding rater perception. The model classifying the presence or absence of tremor achieved the highest performance (accuracy = 0.80, AUROC = 0.82), with TEBR, TEMR, and rCVA as the top contributing features (Figure 7A). In contrast, the model differentiating amplitude constancy from variability showed one of the poorest performances (accuracy = 0.59, AUROC = 0.50), with ApEn, FreqVar, and rCVA emerging as the most influential features (Figure 7B). Rhythm classification also yielded limited performance (accuracy = 0.58, AUROC = 0.70), primarily driven by ApEn, FreqVar, and TEMR. The jerkiness identification model demonstrated modest performance (accuracy = 0.71, AUROC = 0.63), with TEBR, TEMR, and rCVA contributing most strongly. Finally, the frequency classification model achieved good overall accuracy but a lower discriminative ability (accuracy = 0.75, Macro-AUC = 0.49), with sinusoidality, as captured by rSINE, serving as the strongest predictive driver. Overall, the models demonstrated improved fit compared with those reported in.

**Figure 7.**
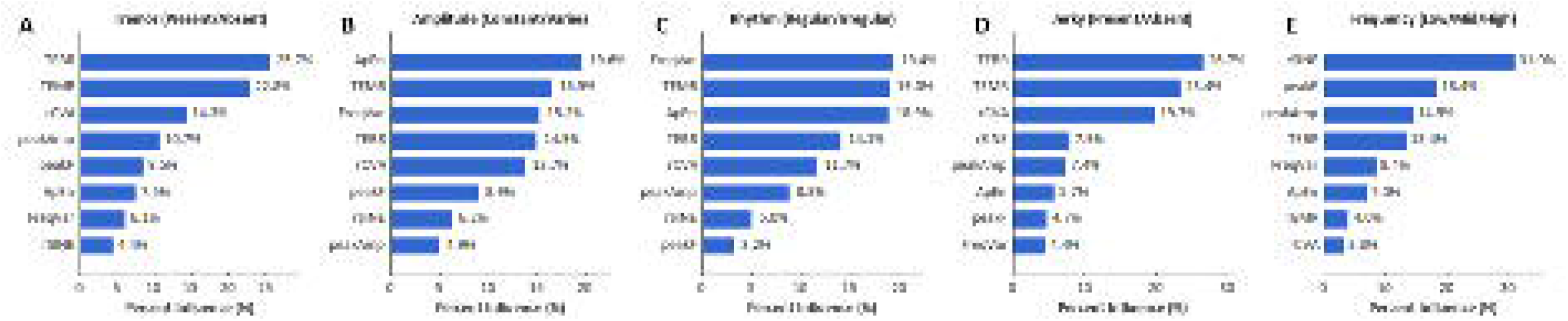
Feature dependence of clinician labels. Percent influence (permutation-based) of each objective feature on classification performance for: (A) Tremor (Present/Absent), (B) Amplitude (Constant/Varies), (C) Rhythm (Regular/Irregular), (D) Jerky (Present/Absent), and (E) Frequency (levels).

## Discussion

Mischaracterization among tremor syndromes remains a significant clinical challenge, particularly when distinguishing ET, PD tremor, and tremor associated with dystonia (Du et al., 2022; Thenganatt and Louis, 2012). Although amplitude and frequency are commonly used quantitative descriptors, they may not fully capture tremor morphology and temporal variability. The goal of this study was to establish a set of objective, intuitive, and easily interpretable metrics for quantifying tremor morphology in a feature space that reflects waveform shape, amplitude stability, and frequency regularity. Unlike prior qualitative classifications, the features defined here: rSINE, rSHAPE, rCVA, rCVD, FreqVar, warping indices (AWI, DWI) and Amplitude Breathing Indices provide simple numerical measures that can be consistently applied across recordings and observers. Each captures a physiologically meaningful aspect of tremor dynamics, ranging from shape regularity and cycle symmetry to amplitude and frequency stability. Although developed here for upper-extremity tremor, the framework is readily extensible to other repetitive movements (e.g., head tremor, nystagmus, stereotypies).

The features rCVD and FreqVar were only weakly correlated. Although both relate to frequency stability, they capture different constructs: rCVD is a time-domain, cycle-to-cycle measure of period jitter that is agnostic to spectral power, whereas FreqVar is a frequency-domain bandwidth that implicitly weights variability by how power is distributed around the peak. Thus, rCVD can be viewed as a “pure” timing-variability metric, while FreqVar reflects a power-weighted frequency spread. Interestingly, rCVD and rSINE were inversely related, indicating that cycle duration changes primarily occur when the waveform deviates from a perfect sinusoid. In other words, loss of sinusoidality accompanies timing irregularity—a plausible reflection of distorted oscillatory drive or competing generators. In our dataset, rSINE and rSHAPE are highly correlated. However, this need not hold universally: for waveforms that are consistent but non-sinusoidal such as sawtooth-like jerk nystagmus (Dell’osso and Daroff, 1975): rSHAPE can remain high (cycle-shape consistency) while rSINE is low (poor sinusoidality), decoupling the two metrics. rSINE value was also inversely proportional to the warping indices validating that any distortion to the cycle shape will lead to reduced sinusoidality. rSINE also varies inversely with the warping indices, indicating that any distortion of cycle shape reduces sinusoidality. Moreover, AWI and DWI are tightly coupled (Figure 5B), suggesting that amplitude warping typically co-occurs with duration warping. Overall rCVA values ranged from (0.04-0.81) were greater than rCVD values that ranged from (0.27-1.28), consistent with prior reports in PD and ET patients where amplitude variability was found to be higher than frequency variability (Gao, 2004). Finally, the thresholds used in “Classification of oscillatory signals by waveform shape and modulation type” are provided as reference points; because features span continua and cluster boundaries are not well-separated, current cutoffs are empirical.

Repetitive oscillations emerge when a phase-locked motor generator in the brain produces a phase-locked movement pattern. This generator may comprise a single loop or multiple interacting oscillatory loops, so cycle-to-cycle changes can reflect their interplay. While neuroimaging and electrophysiology aim to identify these loops, clinical assessment of the resulting symptoms is often limited to qualitative ratings or simple quantitative measures (amplitude, frequency) that may miss key dynamics. Functionally, if a motor circuit is locked into one dominant loop, that loop should be the therapeutic target; if the circuit switches between loops, interventions should vary accordingly. Functionally, if a motor circuit is locked to one dominant loop, that loop should be the therapeutic target; if the circuit switches between loops, treatment should be tailored accordingly. For example, tremor associated with dystonia with a sinusoidal appearance is often attributed to cerebellar pathways, whereas jerky tremor with dystonia is more consistent with basal-ganglia involvement (Erro et al., 2022). This distinction is typically made by clinician judgment. With the Tremor Morphology Toolbox, we provide an objective alternative: the rSINE index quantifies sinusoidality and can separate sinusoidal from jerky morphology reproducibly across recordings and raters, and the treatment should be provided accordingly.

It further suggests that tremor frequency reflects a property of the underlying network, whereas amplitude fluctuations arise from variability in the firing of individual neurons. The Breathing Rate (from the Amplitude Breathing Index) estimates how often the envelope “resets,” i.e., the average rate of the waxing–waning cycles. Lower approximate entropy (e.g., ApEn < 0.3) indicates more regular predictable dynamics, making the breathing pattern more apparent. Together, these features suggest a slower, phase-locked modulatory process that periodically scales the output of the primary oscillation generator. The Breath Modulation Ratio captures the strength (effective gain) of this slower modulation on the oscillation-generating circuitry.

The alignment of clinician perceptions with the newly defined features in the Tremor Morphology Toolkit revealed several notable insights. Amplitude breathing indices had a strong influence on clinicians’ perception of tremor shape, underscoring the complexity of human interpretation and the potential disconnect between descriptive language and the underlying signal dynamics. The model for amplitude stability, despite incorporating these newly defined features, showed poor predictive performance, suggesting inconsistency in how raters perceived and defined this attribute. This highlights a broader ambiguity in the subjective interpretation of amplitude variability across clinicians. The model’s performance may be attributed to two factors. One possibility is that rater perception is inherently more complex than what the current feature set can capture. A more likely explanation is that attributes such as amplitude, jerkiness, and rhythm are interpreted through individualized perceptual heuristics, with each clinician applying unique internal criteria. In contrast, binary judgments such as the presence or absence of tremor show higher perceptual agreement, leading to better model performance. Interestingly, across several perceptual domains—tremor presence, rhythm regularity, and jerkiness—two of the three most influential predictors were derived from envelope-based features rather than cycle-to-cycle measures, suggesting that clinicians may rely more on slow amplitude modulations in the envelope than on fine-grained waveform shape variations when forming perceptual judgments. Frequency classification exhibited good overall accuracy but limited discriminative ability, likely reflecting label imbalance and perceptual ambiguity. The “mid” frequency category was used more frequently by raters, which may have inflated overall accuracy without improving true discriminative power. Furthermore, the absence of clearly defined perceptual boundaries between frequency groups may have introduced uncertainty into the ground truth labels, thereby reducing the model’s AUC performance.

Many studies now apply AI algorithms to classify movement disorders from wearable-sensor data (Brindha et al., 2022; Camara et al., 2015; Dattola et al., 2025; Engin et al., 2007; Ma et al., 2022; Moon et al., 2020; Surangsrirat et al., 2016). While promising, these approaches have two key limitations. First, the “ground truth” is typically defined by clinicians and can therefore be subjective. Second, the derived features are often opaque, reducing clinical interpretability and relevance. These issues highlight the need for intuitive, physically, and physiologically meaningful parameters that will be used in the clinic, as well as research.

Altogether, these metrics offer a descriptive, mechanistically grounded framework for tremor characterization from as little as 30 seconds of data. They bridge qualitative descriptions with quantitative measures that are physiologically interpretable, reproducible, and suitable for cross-subject comparisons, thereby enhancing their translational utility. We encourage researchers to adapt and apply these parameters in conjunction with electrophysiological data to uncover shared mechanisms underlying tremor generation and modulation. Together, such integrative analyses can advance the development of objective, interpretable biomarkers that link behavior to underlying neural dynamics, paving the way for more precise, physiology-informed therapeutic strategies.

## Data Availability

All data produced in the present study are available upon reasonable request to the authors, after appropriate institutional formalities are fulfilled.

## Author’s Roles

- PJ: Writing – original draft, data analysis, Visualization
- YB: Execution, Data Collection, Writing – review & editing
- SP: Investigation, Writing – review & editing
- MV: Investigation, Writing – review & editing
- VF: Writing – review & editing
- MH: Investigation
- HJ: Investigation, Writing – review & editing
- AF: Conceptualization, Methodology, Investigation, Project administration, Funding acquisition, Supervision, Writing – review & editing
- AGS: Conceptualization, Methodology, Investigation, Project administration, Funding acquisition, Supervision, Writing – review & editing

